# BAck Complaints in the Elders - Chiropractic (BACE-C): Design of a cohort study in chiropractic care

**DOI:** 10.1101/19006569

**Authors:** Alan D. Jenks, Trynke Hoekstra, Iben Axén, Katie de Luca, Jonathan Field, Dave Newell, Jan Hartvigsen, Simon D. French, Bart Koes, Maurits W. van Tulder, Sidney M. Rubinstein

## Abstract

**Background:** Low back pain is a common condition among older adults that significantly influences physical function and participation. Compared to their younger counterparts, there is limited information available about the clinical course of low back pain in older people, in particularly those presenting for chiropractic care. Improving our understanding of this patient population and the course of their low back pain may provide input for studies researching safer and more effective care than is currently provided.

**Objectives:** The primary objectives are to examine the clinical course over one year of the intensity, healthcare costs and improvement rates of low back pain in people 55 years and older who visit a chiropractor for a new episode of low back pain.

**Methods:** An international prospective, multi-center cohort study with one-year follow-up. Chiropractic practices are to be recruited in the Netherlands, Sweden, United Kingdom and Australia. Treatment will be left to the discretion of the chiropractor. Inclusion/Exclusion criteria: Patients 55 years and older who are accepted for care having presented to a chiropractor with a new episode of low back pain and who have not been to a chiropractor in the previous six months for an episode of low back pain are to be included, independent of whether or not they have seen another type of health care provider. Patients who are unable to complete the web-based questionnaires because of language restrictions or those with computer literacy restrictions will be excluded as well as those with cognitive disorders. In addition, those with a suspected tumor, fracture, infection or any other potential red flag or condition considered to be a contraindication for chiropractic care will be excluded. Data will be collected using online questionnaires at baseline, and at 2 and 6 weeks and at 3, 6, 9 and 12 months.

**Trial Registration:** Nederlandse Trial Registrar NL7507

## Background

Worldwide, low back pain is the leading cause of years lived with disability and contributes to the global burden of disease (1,2). Low back pain is associated with decreased mobility, reduced social participation, increased isolation and difficulty with activities of daily living and thus has a negative effect on overall health-related quality-of-life in older adults. Older adults with low back pain also more commonly suffer from a range of co-morbidities when compared to those without low back pain (3,4). This results in large costs of care, which are estimated to exceed €400 billion per year worldwide (5).

Low back pain is generally more severe with increasing age (6). For example, one in every four people aged >80 years will report moderate to severe low back pain and people aged >80 years are three times more likely to have high intensity low back pain (scores >50, on a zero to 100 scale) than those aged 50-59 years (7). One-fifth of older adults with low back pain report difficulties in caring for themselves at home or participating in family- and social activities (8). Older people seeking care because of low back pain more commonly receive treatments that have been shown to be ineffective and harmful such as opioid prescription, spinal injections or surgery than younger people seeking care for low back pain (9).

Chiropractors provide a significant portion of care for patients with low back pain (10), and care from chiropractors in the younger and older population appears to be safe and effective (16–18). Unfortunately, existing trials have typically included only younger adults with low back pain, and exclude older adults for various complicating reasons, such as comorbidity and polypharmacy (19-21). As a significant proportion of chiropractors treat older adults (16), it is important to understand the course and characteristics of low back pain in older adults under this care. Perhaps more importantly, chiropractic care may delay functional decline in older adults and improve self-rated health (17,18).

In short, there is a general lack of knowledge regarding low back pain in older adults, but more importantly, data are lacking on course of low back pain for this population in a chiropractic setting (15,19).

The current BACE-C consortium study has been modelled after the ‘BAck Complaints in Elders’ study (BACE), which is an international cohort study devoted to examining back complaints in older people in primary care (23). The BACE-C study is set in chiropractic care. The primary objectives are to examine the clinical course over one year of the intensity, healthcare costs and improvement rates of low back pain in people 55 years and older who visit a chiropractor for a new episode of low back pain.

## Methods

Study design. This study is designed as an international, multi-center prospective cohort study. Data are to be collected from patients 55 and older with low back pain who visit a chiropractor. Follow-up measurements will be scheduled at 2 weeks, 6 weeks, 3 months, 6 months, 9 months and at one year after the first treatment. Participants are to be recruited from the private practices of chiropractors in the Netherlands, Sweden, Australia and the United Kingdom using the same recruitment strategies. The procedures and design outlined in this paper are to be followed by the participating countries and describe a common set of primary outcome measures and patient- and chiropractic factors to be measured. Care will be at the discretion of the participating chiropractors. Ethics approval will be obtained in each participating country prior to data collection.

### Participants

Inclusion criteria: Patients aged 55 and older who consult a chiropractor for an episode of low back pain regardless of duration, either for the first time or patients who have not been to a chiropractor in the previous six months are to be recruited, independent of whether or not they have seen another type of health care provider for the current episode. All low back complaints, with pain in the region from the thoracolumbar 12th rib junction to the first sacral vertebrae, including pelvic pain and pain referral to the leg(s) are to be included.

Exclusion criteria: Patients who are unable to complete the web-based questionnaires because of language restrictions or computer literacy restrictions will be excluded as well as those with cognitive disorders. In addition, those with a suspected tumor, fracture, infection or any other potential red flag or condition considered to be a contraindication for chiropractic care will be excluded.

### Inclusion Procedure

Participating chiropractors will be asked to refer all potential participants who fulfill the inclusion criteria to the online questionnaire, preferably prior to the first appointment. Participants will be briefly informed about the study procedures over the phone when they call to make an appointment or during the initial consultation with the chiropractor. The chiropractor or chiropractic assistant will ask for the patient’s permission to send an email with a link to the informed consent and baseline questionnaire, so that it can be completed at home prior to the first visit or as soon as possible and no later than two weeks after the initial visit. Figure 1 shows the proposed flow of patient inclusion.

**Figure 1.**
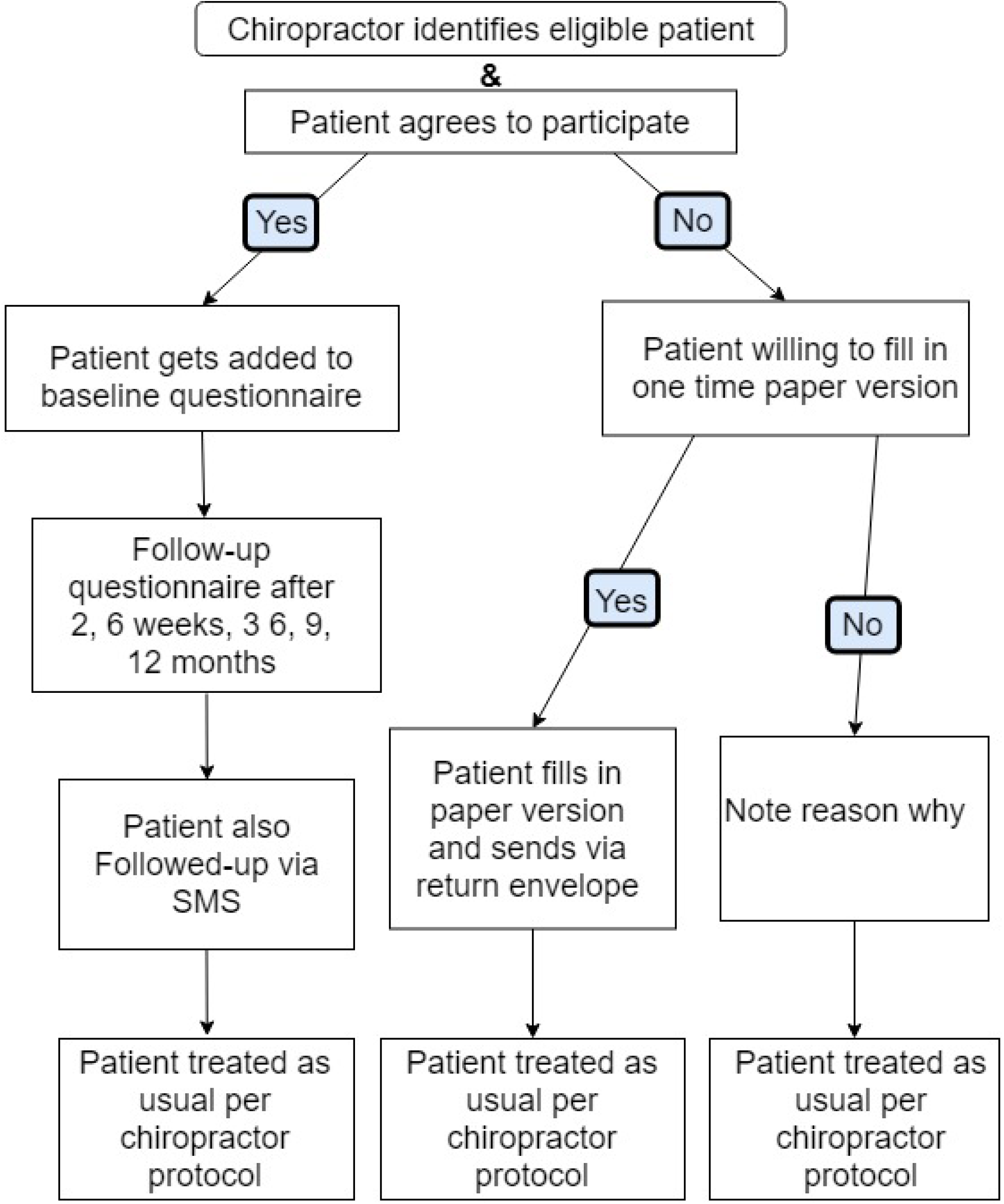
Flow Chart of BACE-C study.

### Questionnaires

Links to the questionnaires will be sent by email and completed as a web-based questionnaire at baseline, 2 and 6 weeks, and at 3, 6, 9 and 12 months after the initial visit. In Sweden data will not be collected after 6 weeks because of logistical burden. Table 1 shows the measurements per follow-up round and the time frame for data collection.

**Table 1:**
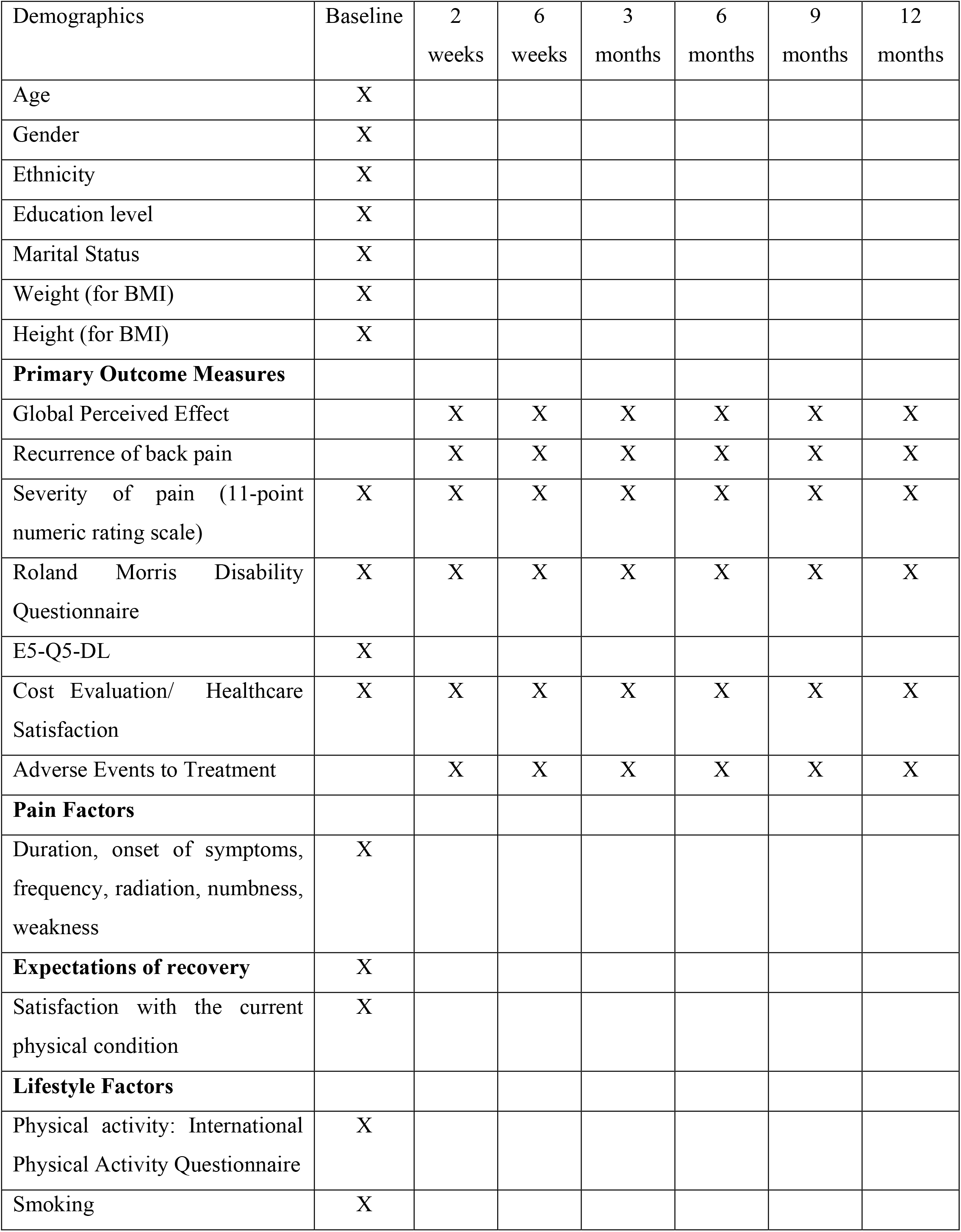
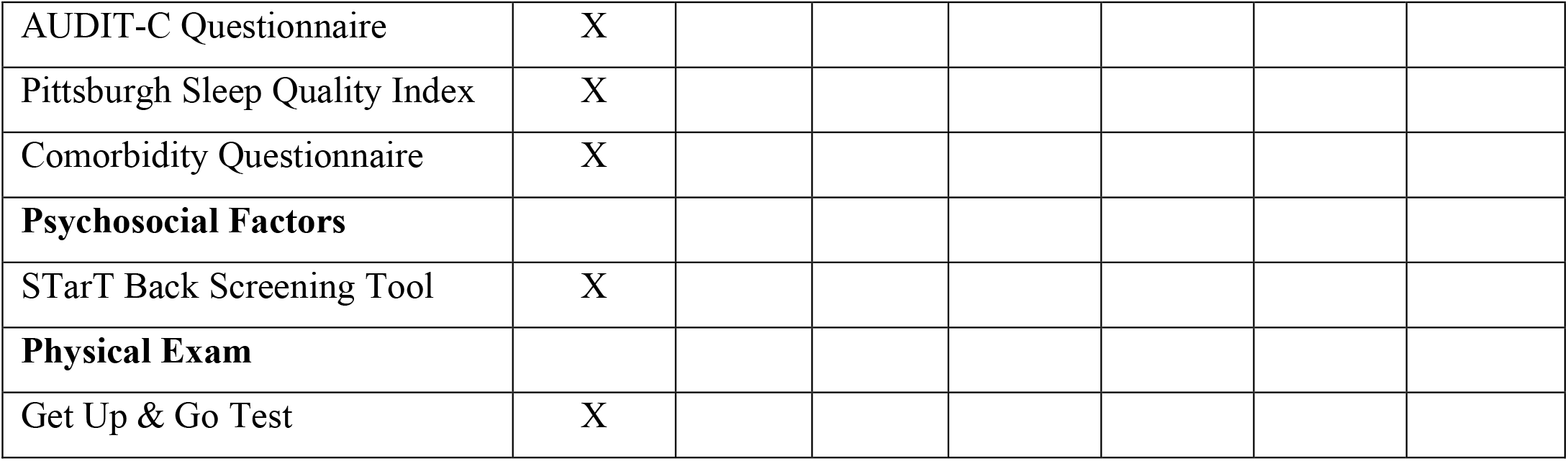
Content of the patient questionnaires.

#### The primary outcome measures are

1) low back pain intensity, 2) back-specific functional status and 3) global perceived effect. As a secondary measure, 4) healthcare costs will be measured. All outcomes are self-reported.

#### Patient-related factors

The following factors will be measured at baseline: 1) sociodemographic characteristics (i.e. age, gender, marital status, education level, height, weight); 2) physical activity (measured with the International Physical Activity questionnaire (20)); 3) other lifestyle variables smoking; measured by pack years (21), alcohol use measured by the short version of the AUDIT-C (22,23), sleeping habits; measured by the short version of the Pittsburgh Sleep Quality Index (24); 4) comorbidities using the Self-administered Comorbidity Questionnaire (25) and 7) indicator screening tool (STarT Back) for poor outcome (26) and 5) quality-of-life measured with the EQ-5D-5L at baseline only. In Sweden the EQ-5D-3L will be used. The EQ-5D measures five dimensions: mobility, self-care, usual activities, pain/discomfort and anxiety/depression (28,29).

In the Netherlands, each chiropractor will also perform at the first consult the “timed Up & Go” test (30). The “timed Up & Go” test is composed of a variety of movements which are necessary for daily activities: walking, standing up, turning, stopping, and sitting down; and predictive of falls in the elderly (30). In previous studies, this test showed associations with quality-of-life scores (31).

#### Pain

Pain intensity will first be measured using an 11-point numerical rating scale (NRS) (32) in which 0 represents ‘no pain ‘and 10 represents ‘the worst pain ever’. Second, several questions about the severity and reoccurrence of complaints will be asked at all follow-up measurements (table 1).

#### Back-specific functional status

Functional status will be measured at baseline and all follow-up intervals using the Roland Morris Disability Questionnaire (RMDQ) (33), in which total score can range from 0 (no disabilities) to 24 (severe disabilities).

#### Global perceived effect

Global perceived effect (GPE) will be measured on a 7-point scale, ranging from ‘completely recovered’ to ‘worse than ever’ (33,34). Patients will be asked to provide additional (open-ended) explanation if they report worse or much worse global perceived effect compared to the previous follow-up measurement. GPE will be dichotomized for the analyses as follows: ‘completely recovered’ and ‘much better’ will be considered ‘improved’, while all other responses will be considered ‘not improved’ (35).

#### Healthcare consumption

Healthcare consumption will include the use of all primary health care (e.g. general practitioner, physiotherapist), all secondary healthcare (e.g. hospital based neurologist, orthopedic surgeon), hospitalization, complementary care (e.g. acupuncture, dry needling, massage) as well as the use of both prescribed and over the counter medication. Questions were adapted based on the iMTA medical consumption questionnaire (36). Healthcare consumption characteristics will be valued in accordance with costing guidelines of each participating country, such as the Dutch Manual of Costing (37).

#### Chiropractor-related factors

These variables will be obtained from the chiropractors themselves: 1) sociodemographic (age, gender), school attended (school, year of graduation), and types of treatments commonly delivered in their practice.

In the Netherlands and in Sweden, each chiropractor will be asked to fill in several questions about their expectations of patient recovery. This will be asked at the first four treatment visits.

### Statistical analyses

Descriptive analyses: Baseline variables will be presented as percentages for categorical variables and as means plus standard deviations for continuous variables. In case of non-normal distributions, continuous variables will be described as medians with corresponding interquartile ranges. Furthermore, descriptive information of the primary and secondary outcome variables will be presented for baseline and all follow-up intervals. Descriptive analyses will be conducted for the entire data set from all participating countries as well as stratified for each country.

The primary objective will be answered using the entire data set from all participating countries and subsequently stratified by country.

In addition, the primary objective will be answered for each primary outcome separately by multilevel models with three levels (observations over time clustered within patients, clustered within practices). Country will be included as a covariate in the models (as dummy variables) (38). The models will thus include time as a continuous variable as well as country as independent variables. Potential need for time squared and time cubed will be investigated by assessing the significance level of the quadratic and/or cubic terms. A random intercept will be included a priori. The need for a random slope for time will be investigated by the likelihood ratio test, in a stepwise manner (38).

The clinical course of pain and back-specific functional status will be analyzed by linear multilevel models, global perceived effect by logistic multilevel models and healthcare costs by a linear multilevel model with bootstrapped confidence intervals because of the expected skewed distribution of the cost data. We will report regression coefficients (linear models), odds ratios (logistic models), corresponding 95% confidence intervals and two-sided p-values.

## Discussion

This study is to our knowledge the first large-scale, prospective, multicenter, international study to be conducted in a chiropractic setting and the first one focusing on older adults with low back pain consulting a chiropractor. The primary objectives of the BACE-C study are to examine the clinical course over one year of the intensity, healthcare costs and improvement rates of low back pain in people 55 and older who visit a chiropractor for a new episode of low back pain. By understanding the impacts of various factors on the course and treatment of low back pain in the elderly population, this large data set will allow us to provide input for the development of future feasibility intervention studies in this patient group. We invite other research groups worldwide to join the BACE-C consortium.

### Data Management, Storage and Security

Data will be stored on institutional network drives with firewalls and security measures in place according to national and European Union data protection regulations. Hard copy records will be stored in a locked cabinet in a secure location. Access to records and data will be limited to study personnel. Study data will be de-identified and a master log file with identifiers will be kept and stored separately from the data. Only anonymized data will be used for analyses.

## Data Availability

Data sharing is not applicable to this article as no datasets were generated or analyzed during the current study.

## Declarations

### Ethics approval and consent to participate

This protocol has received ethical approval from the Medical Ethics Committee of the Vrije University Medical Center number, the Netherlands ethics number 2017-618, the Karolinska Institutet, Sweden ethics number 2018/474-31/2 and the Science & Engineering Subcommittee, Human Research Ethics Committee at Macquarie University, Australia, Project ID 5460. Currently the UK are awaiting ethics approval.

### Consent for publication

Written informed consent was obtained from the patient for publication of their individual details and accompanying images in this manuscript. The consent form is held by the authors’ institution in a secured server and is available for review by the Editor-in-Chief.

### Authors’ contributions

Concept and design: Alan Jenks, Trynke Hoekstra, Bart Koes, Maurits van Tulder, Sidney Rubinstein Drafting of this manuscript: Alan Jenks, Trynke Hoekstra, Maurits van Tulder, Sidney Rubinstein Critical revision of the article for important intellectual content: All members Final approval of the article: All members

## Acknowledgements

We would like to thank all the original authors of the BACE consortium in the Netherlands, Brazil and Australia for their input and suggestions. Additionally, we thank Jen Walraven, Mojdeh Khorrami and Mariana Fadil Romao for their comments on drafts of this manuscript.

## References

1. Hoy D, March L, Brooks P, Blyth F, Woolf A, Bain C, et al. The global burden of low back pain: Estimates from the Global Burden of Disease 2010 study. Ann Rheum Dis. 2014;73(6):968–74.

2. Buchbinder R, van Tulder M, Öberg B, Costa LM, Woolf A, Schoene M, et al. Low back pain: a call for action. Vol. 391, The Lancet. 2018.

3. Cedraschi C, Luthy C, Allaz A-F, Herrmann F, Ludwig C. The impact of low back pain on health-related quality of life in seniors living in the community. Eur spine journal Conf Annu Congr EUROSPINE 2017 Irel [Internet]. 2017;26(2 Supplement 1):S305. Available from: http://onlinelibrary.wiley.com/o/cochrane/clcentral/articles/767/CN-01421767/frame.html

4. Hartvigsen J, Frederiksen H, Christensen K. Back and neck pain in seniors - Prevalence and impact. Eur Spine J. 2006;15(6):802–6.

5. Manogharan S, Kongsted A, Ferreira ML, Hancock MJ. Do older adults with chronic low back pain differ from younger adults in regards to baseline characteristics and prognosis? Eur J Pain (United Kingdom). 2017;21(5):866–73.

6. Walker BF, Muller R, Grant WD. Low back pain in Australian adults. Health provider utilization and care seeking. J Manipulative Physiol Ther. 2004;27(5):327–35.

7. Williams JS, Ng N, Peltzer K, Yawson A, Biritwum R, Maximova T, et al. Risk factors and disability associated with low back pain in older adults in low- and middle-income countries. Results from the WHO study on global AGEing and adult health (SAGE). PLoS One. 2015;10(6):1–21.

8. Rudy TE, Weiner DK, Lieber SJ, Slaboda J, Boston JR. The impact of chronic low back pain on older adults: A comparative study of patients and controls. Pain. 2007;131(3):293–301.

9. Hartvigsen J, Hancock MJ, Kongsted A, Louw Q, Ferreira ML, Genevay S, et al. What low back pain is and why we need to pay attention. Lancet. 2018;391(10137):2356–67.

10. Beliveau PJH, Wong JJ, Sutton DA, Simon N Ben, Bussières AE, Mior SA, et al. The chiropractic profession: A scoping review of utilization rates, reasons for seeking care, patient profiles, and care provided. Chiropr Man Ther. 2017;25(1):1–17.

11. Hebert JJ, Stomski NJ, French SD, Rubinstein SM. Serious Adverse Events and Spinal Manipulative Therapy of the Low Back Region: A Systematic Review of Cases. J Manipulative Physiol Ther [Internet]. 2015;38(9):677–91. Available from: http://dx.doi.org/10.1016/j.jmpt.2013.05.009

12. Rubinstein SM, Terwee CB, Assendelft WJJ, de Boer MR, van Tulder MW. Spinal Manipulative Therapy for Acute Low Back Pain. Spine (Phila Pa 1976) [Internet]. 2013;38(3):E158–77. Available from: http://content.wkhealth.com/linkback/openurl?sid=WKPTLP:landingpage&AN=00007632-201302010-00020

13. Rubinstein SM, Middelkoop M Van, Boer MR De, Tulder MW Van, van Middelkoop M, Assendelft WJ, et al. Spinal Manipulative Therapy for Chronic. Spine (Phila Pa 1976) [Internet]. 2011;36(2):2–5. Available from: http://www.ncbi.nlm.nih.gov/pubmed/21328304 %5Cn http://onlinelibrary.wiley.com/doi/10.1002/14651858.CD008112.pub2/pdf/standard

14. Vos T, Flaxman AD, Naghavi M, Lozano R, Michaud C, Ezzati M, et al. Years lived with disability (YLDs) for 1160 sequelae of 289 diseases and injuries 1990-2010: A systematic analysis for the Global Burden of Disease Study 2010. Lancet. 2012;380(9859):2163–96.

15. de Luca KE, Fang SH, Ong J, Shin KS, Woods S, Tuchin PJ. The Effectiveness and Safety of Manual Therapy on Pain and Disability in Older Persons With Chronic Low Back Pain: A Systematic Review. J Manipulative Physiol Ther [Internet]. 2017;40(7):527–34. Available from: https://doi.org/10.1016/j.jmpt.2017.06.008

16. Moore C, Swain M, Luca K De, Hartvigsen J, Wong AYL. Characteristics of chiropractors who manage people aged 65 and older : A nationally representative sample of 1903 chiropractors. 2019;(February):1–9.

17. Weigel P, Hockenberry JM, Bentler SE, Obrizan M, Kaskie B, Jones MP, et al. A longitudinal study of chiropractic use among older adults in the United States. Chiropr Osteopat [Internet]. 2010;18(1):34. Available from: http://chiromt.biomedcentral.com/articles/10.1186/1746-1340-18-34

18. Weigel PAM, Hockenberry JM, Wolinsky FD. Chiropractic use in the medicare population: Prevalence, patterns, and associations with 1-year changes in health and satisfaction with care. J Manipulative Physiol Ther. 2014;37(8):542–51.

19. Goertz CM., Salsbury SA., Vining RD., Long CR., Andresen AA., Jones ME., et al. Collaborative Care for Older Adults with low back pain by family medicine physicians and doctors of chiropractic (COCOA): Study protocol for a randomized controlled trial. Trials [Internet]. 2013;14(1):1–18. Available from: http://www.scopus.com/inward/record.url?eid=2-s2.0-84872196848&partnerID=40&md5=cd0b06c1cf5da88d346f6c99fa6cbf30

20. Carvalho FA, Morelhão PK, Franco MR, Maher CG, Smeets RJEM, Oliveira CB, et al. Reliability and validity of two multidimensional self-reported physical activity questionnaires in people with chronic low back pain. Musculoskelet Sci Pract. 2017;27:65–70.

21. The Association Between Cigarette Smoking and Back Pain in Adults.

22. Weitkunat R, Coggins CRE, Sponsiello-Wang Z, Kallischnigg G, Dempsey R. Assessment of cigarette smoking in epidemiologic studies. Beitrage zur Tab Int Contrib to Tob Res. 2013;25(7):638–48.

23. Frank D, DeBenedetti AF, Volk RJ, Williams EC, Kivlahan DR, Bradley KA. Effectiveness of the AUDIT-C as a screening test for alcohol misuse in three race/ethnic groups. J Gen Intern Med. 2008;23(6):781–7.

24. Buysse DJ, Charles F, Reynolds III, Monk T, Berman S, Kupfer D. The Pittsburgh Sleep Quality Index: A new Instrument for Psychiatric Practic and Research. Psychiatry Res [Internet]. 1988;(28):193–213. Available from: http://www.ncbi.nlm.nih.gov/pmc/articles/PMC3023245/ %0A http://dx.doi.org/10.1016/S0735-1097(01)01746-6 %0A http://eutils.ncbi.nlm.nih.gov/entrez/eutils/elink.fcgi?dbfrom=pubmed%7B%7Did=2748771%7B%7Dretmode=ref%7B%7Dcmd=prlinks %0A http://dx.doi.org/10.1016

25. Sangha O, Stucki G, Liang MH, Fossel AH, Katz JN. The self-administered comorbidity questionnaire: A new method to assess comorbidity for clinical and health services research. Arthritis Rheum. 2003;49(2):156–63.

26. Jasper D. Bier, Raymond W.J.G. Ostelo, Miranda L. van Hooff, Bart W. Koes APV. Validity and Reproducibility of the STarT Back Tool (Dutch Version) in Patients With Low Back Pain in Primary Care Settings. 2017;97(6):659–68.

27. Greenspan SL, Bouxsein ML, Melton ME, Kolodny a H, Clair JH, Delucca PT, et al. Precision and discriminatory ability of calcaneal bone assessment technologies. J Bone Miner Res [Internet]. 1997;12(8):1303–13. Available from: http://www.ncbi.nlm.nih.gov/pubmed/9258762

28. Mangen MJJ, Bolkenbaas M, Huijts SM, van Werkhoven CH, Bonten MJM, de Wit GA. Quality of life in community-dwelling Dutch elderly measured by EQ-5D-3L. Health Qual Life Outcomes [Internet]. 2017;15(1):1–6. Available from: http://dx.doi.org/10.1186/s12955-016-0577-5

29. Whynes DK, McCahon RA, Ravenscroft A, Hodgkinson V, Evley R, Hardman JG. Responsiveness of the EQ-5D health-related quality-of-life instrument in assessing low back pain. Value Heal [Internet]. 2013;16(1):124–32. Available from: http://dx.doi.org/10.1016/j.jval.2012.09.003

30. Podsiadlo D, Richardson S. The Timed “Up & Go”: A Test of Basic Functional Mobility for Frail Elderly Persons. J Am Geriatr Soc [Internet]. 1991;39(2):142–8. Available from: http://doi.wiley.com/10.1111/j.1532-5415.1991.tb01616.x

31. Dobson F. Timed Up and Go test in musculoskeletal conditions. J Physiother [Internet]. 2015; 61(1):47. Available from: http://dx.doi.org/10.1016/j.jphys.2014.11.003

32. Von Korff M, Jensen MP, Karoly P. Assessing global pain severity by self-report in clinical and health services research. Spine (Phila Pa 1976). 2000;25(24):3140–51.

33. Chiarotto A, Boers M, Deyo RA, Buchbinder R, Corbin TP, Costa LOP, et al. Core outcome measurement instruments for clinical trials in nonspecific low back pain. Pain [Internet]. 2018;159(3):1. Available from: http://insights.ovid.com/crossref?an=00006396-900000000-99091

34. Luijsterburg PA, Verhagen AP, Ostelo RW, van den Hoogen HJ, Peul WC, Avezaat CJ, et al. Conservative treatment in patients with an acute lumbosacral radicular syndrome: design of a randomised clinical trial [ISRCTN68857256]. BMC Musculoskelet Disord [Internet]. 2004;5(1):39. Available from: http://bmcmusculoskeletdisord.biomedcentral.com/articles/10.1186/1471-2474-5-39

35. Scheele J, Luijsterburg PA, Ferreira ML, Maher CG, Pereira L, Peul WC, et al. Back complaints in the elders (BACE); design of cohort studies in primary care: an international consortium. BMC Musculoskelet Disord [Internet]. 2011;12:193. Available from: http://www.ncbi.nlm.nih.gov/pubmed/21854620

36. Bouwmans C, Krol M, Severens H, Koopmanschap M, Brouwer W, Roijen LH Van. The iMTA Productivity Cost Questionnaire: A Standardized Instrument for Measuring and Valuing Health-Related Productivity Losses. Value Heal [Internet]. 2015;18(6):753–8. Available from: http://dx.doi.org/10.1016/j.jval.2015.05.009

37. Kanters TA, Bouwmans CAM, Van Der Linden N, Tan SS, Hakkaart-van Roijen L. Update of the Dutch manual for costing studies in health care. PLoS One. 2017;12(11):1–11.

38. Twisk JWR. Applied Mixed Model Analysis [Internet]. Cambridge University Press; 2019. Available from: https://www.cambridge.org/core/product/identifier/9781108635660/type/book

